# Mandatory public health measures for COVID-19 are associated with improved mortality, equity and economic outcomes

**DOI:** 10.1101/2021.02.11.21251580

**Authors:** Brita E. Lundberg, Kathyrn McDonald

**Affiliations:** Lundberg Health Advocates, 1180 Beacon St., #4B, Brookline, MA 02446; Bloomberg Distinguished Professor at Johns Hopkins Bloomberg School of Public Health, Johns Hopkins Carey Business School and Schools of Medicine and Nursing

## Abstract

We review approaches to the COVID-19 pandemic in a systematic way by comparing countries/states representative of the mandatory vs. voluntary approach to non-pharmaceutical interventions in Europe and the US. We use a comparative tabular format to examine differences in mortality, economic impact and equity between regions with mandatory versus voluntary policies. Mandatory shelter-in-place policies were associated with 3 to 4 fold lower population adjusted mortality in the US model and 11 fold lower in the European one. We conclude that voluntary policies are less effective, based on historical precedent and the current analysis. Moreover, effects on health equity mirrored the increased mortality outcomes of voluntary policies and there was no apparent economic benefit associated with voluntary measures.

## Introduction

Public health interventions to address the COVID-19 pandemic including shelter-in-place, quarantine, isolation and travel restrictions, have varied from no policy to mandatory enforcement. We studied the relationship between mandatory compared to voluntary enforcement of shelter-in-place policies and COVID-19 mortality, equity and economic outcomes.

## Methods

We analyzed mortality data from regions with similar demographics and comprehensive health data but markedly contrasting public health policies. In the US, we compared Colorado and Washington, which implemented early mandatory shelter-in-place orders, and Massachusetts, which issued this measure as a voluntary health advisory. In Europe, we compared Norway’s mandatory shelter-in-place policy to Sweden’s voluntary one.

## Results

We found differences in the number of cases and mortality rates in the two groups (**Table**). Demographic characteristics known to be risk factors for COVID-19, including age, obesity, lower socioeconomic status (as measured by Gini coefficient), and percent nonwhite, were not statistically different between the comparison groups with the exception of density. In the United States, higher population density is associated with decreased mortality from COVID-19.^1^ Yet population-dense Massachusetts experienced a higher case burden and mortality than either Colorado or Washington, with a 4.7-fold higher population-adjusted mortality rate than Washington and 3.7-fold higher than Colorado (p<0.0001; **Figure**). Sweden recorded a population-adjusted mortality rate 11.4-fold higher than Norway (p<0.0001). Mortality among non-Whites was also worse in Massachusetts compared to the two States with stricter public health measures (p<0.0001).

### Economic impacts

Economic impact as measured by GDP was not markedly different between regions that practiced mandatory public health measures compared to those implementing voluntary ones (Table). Sweden’s GDP fell 8.6% in the second quarter, compared to Norway’s 7.4% [Table]; somewhat greater losses in GDP contribution were observed in Massachusetts compared to Colorado or Washington. As measured by VSL (value of a statistical life), the $97.5 billion economic cost to Massachusetts was significantly higher than the $22.6 and $21.8 billion recorded by Washington and Colorado respectively. This difference was pronounced for Sweden, whose economic losses as measured by VSL were 21.3 times that of Norway.

## Discussion

Shelter-in-place has been shown to be essential to containing the COVID-19 epidemic^2^ and mandatory isolation and quarantine key to halting its exponential growth.^3^ In the influenza epidemic of 1918, adoption of mandatory public health control measures at the beginning of the epidemic and resumption of those measures when cases began to rise resulted in lower mortality rates.^4^ In the COVID-19 pandemic, our results show that adoption of mandatory public health shelter-in-place orders resulted in lower mortality rates and a fewer deaths both in the United States and in Europe.

### Other restrictions

In the US, isolation and quarantine of known/exposed cases were mandatory only in the first month of the pandemic, and only for returning travelers.^5^ Mandatory travel restrictions were imposed in Norway and in some parts of Colorado; however, it is plausible that travel restrictions imposed by neighboring jurisdictions may effectively have further locked down Sweden and Washington State and likely contributed to improved mortality outcomes compared to the expected trajectory from internal policy actions. Norway’s substantial success may be further attributed to the mandatory enforcement of all four public health policies: shelter-in-place, quarantine or isolation of exposed / known cases, and travel restrictions.

### Economic benefits

The benefits in the US from social distancing were estimated at the beginning of the pandemic to outweigh the costs to gross domestic product (GDP) by roughly $5.2 trillion,^6^ yet preserving economic health has been used as a justification to refrain from mandatory public health policies. Our results show that both Sweden and Massachusetts adopted voluntary shelter-in-place measures yet experienced far greater VSL losses, especially, in Massachusetts, among minority populations, and somewhat greater losses to GDP.

We conclude that mandatory enforcement of public health measures in response to COVID-19 is likely a critical contributor to decreased mortality and attenuated economic impact of the epidemic. When case numbers rise, mandatory measures should be reinstated in an equitable, thoughtful manner that promotes public engagement and supports livelihoods in order to limit viral spread, prevent death, decrease inequities and preserve economic health.

## Limitations

Study limitations include reliance on provisional data and assumptions applied to the model.

## Data Availability

All data, unless otherwise specified, is extracted from the Johns Hopkins COVID data map: https://coronavirus.jhu.edu/map.html; and from relevant federal, state, and country public health websites.

## Conflicts of interest: None declared

There are no financial or other relationships that might lead to a conflict of interest.

## Key points

- Mandatory enforcement of public health measures in response to COVID-19 is likely a critical contributor to decreased mortality and attenuated economic impact of the epidemic.
- Mandatory measures should be adopted in an equitable, thoughtful manner that promotes public engagement and supports livelihoods in order to limit viral spread, prevent death, decrease inequities and preserve economic health.

---

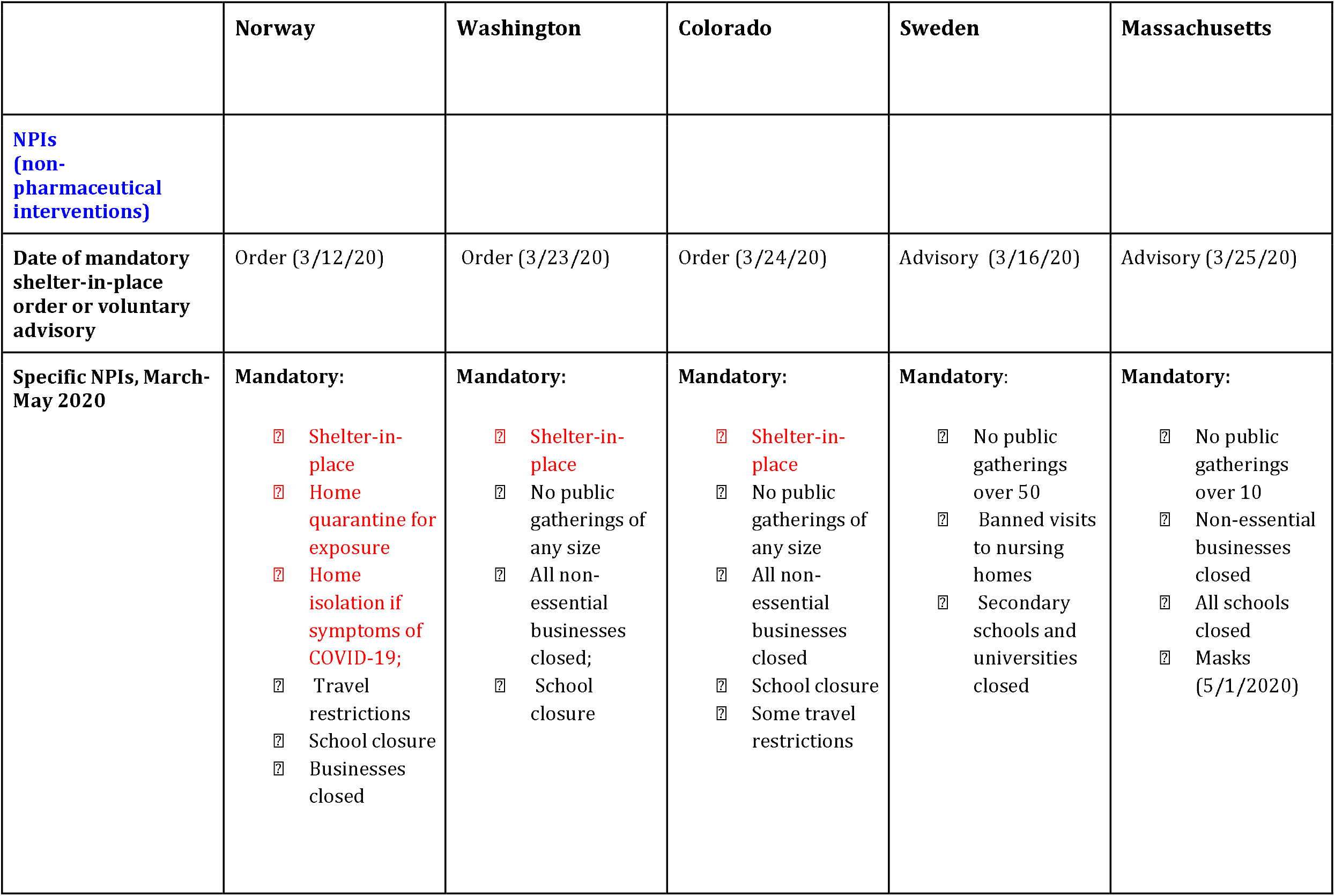
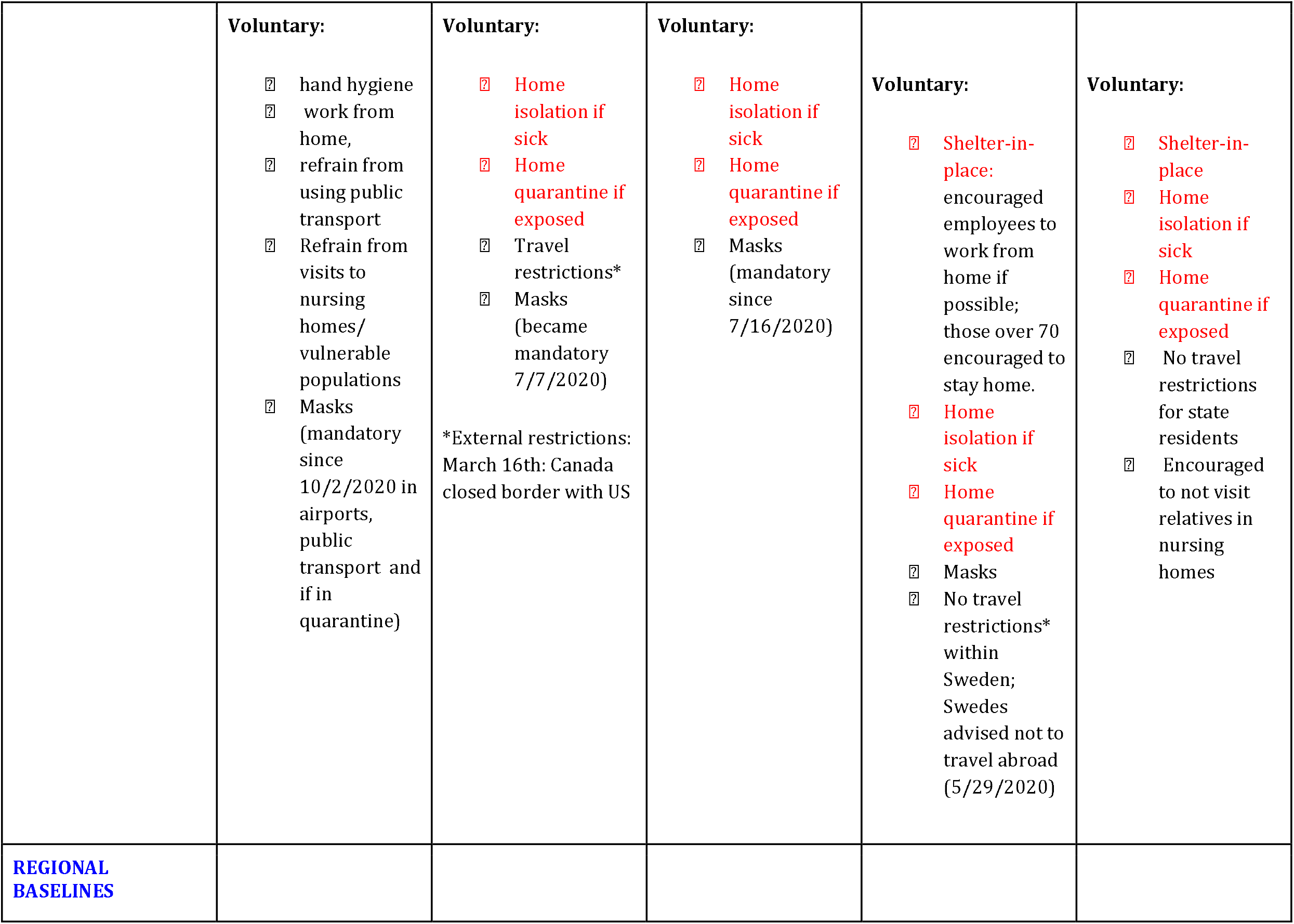
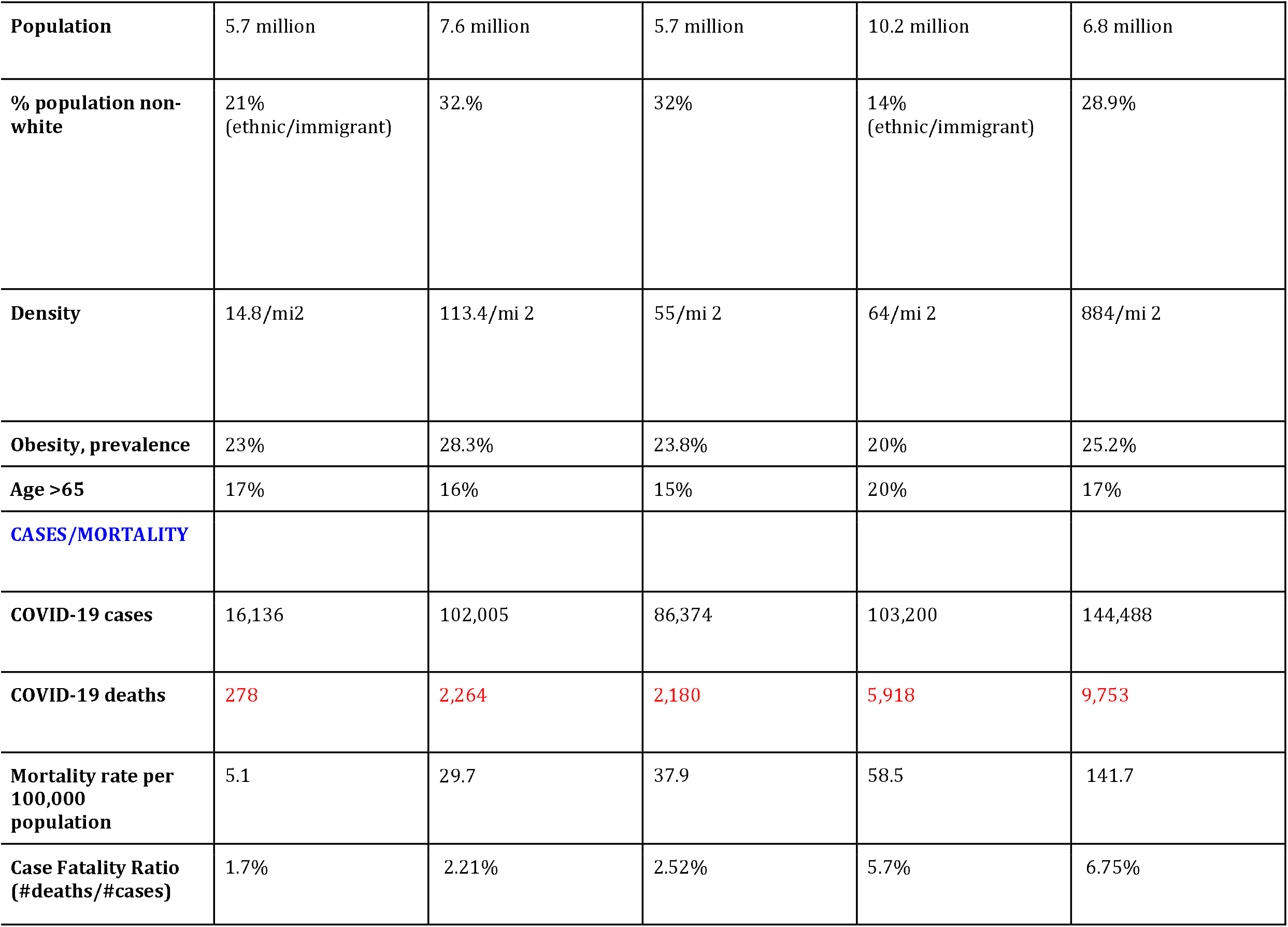
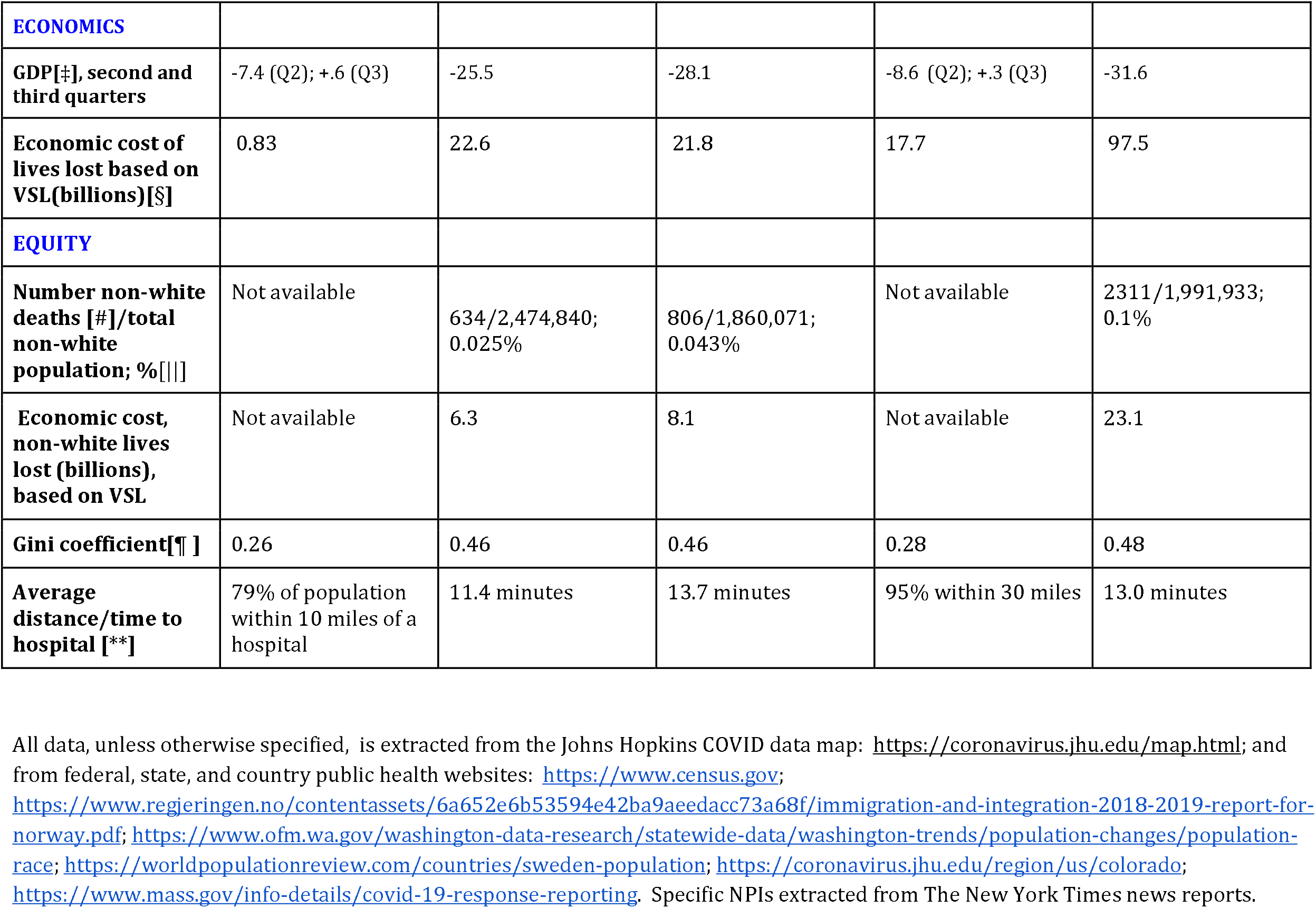
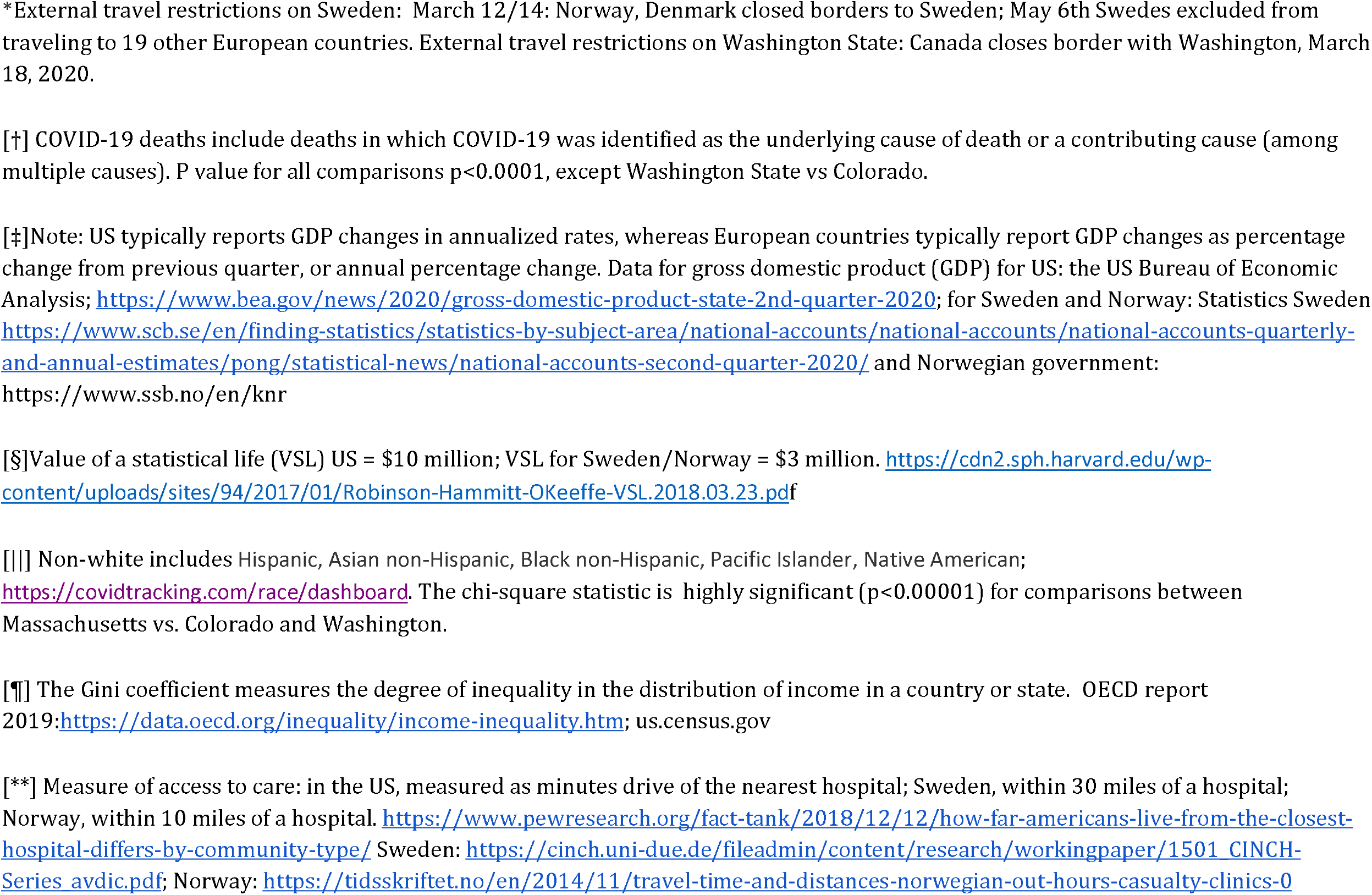
Differences between states and countries implementing voluntary vs. mandatory NPIs

