## Supplemental figure for "Mandatory public health measures for COVID-19 are associated with improved mortality, equity and economic outcomes"

**Figure legend:** Mortality rate per 100,000 from March through November 1, 2020 in Selected States and Countries.

The figure plots mortality in selected states and countries with contrasting voluntary vs. mandatory shelter-in-place policies. Time vs. mortality rate/100,000, normalized for population. Data extracted from: Johns Hopkins COVID data map: <https://coronavirus.jhu.edu/map.html>

**Red arrow:** indicates date of Norwegian shelter-in-place order (March 12th) and travel restrictions imposed on Sweden by Norway and Denmark (March 12th).

**Blue arrow:** indicates Sweden’s voluntary shelter-in-place order (March 16th) and travel restrictions placed by Canada on Washington State.

**Not shown**: Washington and Colorado enact shelter-in-place orders March 23 and 24; Massachusetts enacts voluntary shelter-in-place March 25, 2020.

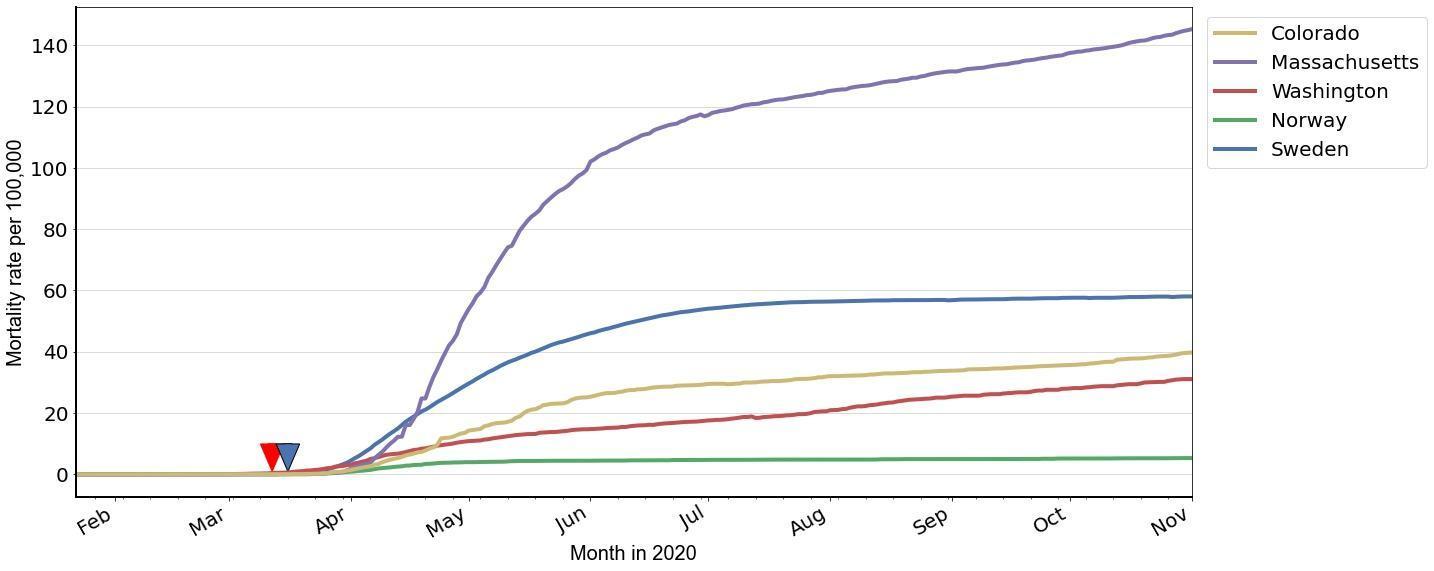
